# Ultra–short-wave diathermy shortens the course of moderate and severe COVID-19: a randomized trial

**DOI:** 10.1101/2021.01.28.21250163

**Authors:** Liangjiang Huang, Qian Li, Shah Zulfiqar Ali Sayed, Nasb Mohammad, Bin Chen, MPhil Ali Iftikhar, Lingfeng Xie, Jifa Hu, Hong Chen

## Abstract

**Question:** Is ultra-short-wave diathermy (USWD) safe and effective in coronavirus disease 2019 (COVID-19) ?

**Design:** Single-centre, evaluator-blinded, two-arm, parallel design, randomized controlled clinical trial.

**Participants:** Moderate and severe COVID-19 patients with acute respiratory syndrome.

**Intervention:** USWD for 10 minutes twice daily for 12 consecutive days along with standard medical treatment (USWD group, n = 25), versus standard medical treatment alone (control group, n = 25).

**Outcome measures:** The primary outcomes were the duration of recovery and negative conversion rate of severe acute respiratory syndrome coronavirus 2 (SARS-CoV-2) on days 7, 14, 21, and 28. Secondary outcomes included clinical status (seven-category ordinal and systemic inflammatory response syndrome (SIRS) scores), computed tomography (CT), routine blood tests, and adverse events.

**Results:** Time to clinical recovery (USWD 36.84±9.93 vs. control 43.56±12.15, P = 0.037) was significantly shortened with a between-group difference of 6.72 days. Clinical status was improved with significant between-group differences on day 28 (SIRS, P = 0.011; seven-category scale, P = 0.003). The rate of RNA negative conversion at days 7 (P = 0.066), 14 (P = 0.239), 21 (P = 0.269), and 28 (P = 0.490) was statistically insignificant. Moreover, insignificant differences were observed in the artificial intelligence-assisted CT analysis. No treatment-associated adverse events or worsening of pulmonary fibrosis were observed.

**Conclusion:** USWD, as adjunctive therapy, shortened the recovery course and improved clinical status of patients with COVID-19 without aggravating pulmonary fibrosis. the findings are limited due to the small sample size and early termination.

**Registration:** ChiCTR2000029972

## Introduction

The outbreak of the coronavirus disease 2019 (COVID-19) pandemic has prompted efforts to manage threat to the well-being of populations worldwide.^1,2,3,4^ The second wave of the COVID-19 epidemic has emerged in some countries^5^; however, to date, no effective treatment has been confirmed. In response to the critical demand for high-quality clinical guidance at the peak of the outbreak in China, guidelines were published to clarify that physical therapy could play an important role in managing COVID-19.^6,7,8,9^ Moreover, it was recommended that supportive therapies such as ultra–short-wave diathermy (USWD) could boost the immune responses and inhibit inflammation.^10,11,12,13^

USWD has been used in the field of physical therapy and rehabilitation for many decades ^14,15,16^, but the evidence for its application as part of the COVID-19 management is debatable.^10^ During the outbreak of severe acute respiratory syndrome (SARS), USWD was widely employed by rehabilitation professionals in China to reduce pneumonia inflammation. Zhang et al. evaluated the efficacy of both USWD and conventional therapy in 38 patients with SARS, where USWD was used as an adjuvant treatment in addition to standard therapy. They concluded that USWD could accelerate recovery and reduce the length of hospital stay.^17^

The pathogens causing COVID-19 and SARS are both coronaviruses, but the clinical manifestation, death rate, and pathological changes, especially fibrosis, are different. Moreover, high-quality evidence to recommend the application of USWD in improving the pulmonary conditions is still lacking, and there is a complete absence of evidence on the use of USWD in treating COVID-19 due to its novel nature. The scant evidence for the safety and efficacy of USWD makes its application in clinical settings questionable. Therefore, we designed a randomized controlled trial to investigate the clinical efficacy and safety of USWD in managing COVID-19.

## Method

### design

This single-centre, evaluator-blinded, two-arm (1:1 ratio) parallel design, superiority randomized controlled trial, and prospectively registered on 17 February 2020 with the Chinese Clinical Trials Registry (Identifier: ChiCTR2000029972). The study was conducted in accordance with the relevant regulations and guidelines of good clinical practice and the Declaration of Helsinki. Patient recruitment, randomization, and study events are visually described in the CONSORT flow diagram (Figure 1). Participants were recruited between 18 February 2020 and 20 April 2020. Before randomization, written and verbal informed consent was obtained and an informative essay that clearly showed the risks and the supposed benefits accompanying the participation were provided to each patient.A statistician, who was not part of the study, created an online randomization plan on www.randomization.com using the permuted blocks method with small blocks of various sizes. This was an assessor-blinded, controlled study, and because of the nature of the interventions, it could not be therapist- or patient-blinded; however, a well-trained health care team comprising two evaluators, two statisticians, and two data collectors were blinded to the groups/treatment allocation. The outcomes were independently documented based on a mutual consensus between the data collectors (Figure 1).The data collection forms developed for this trial consisted of medical history forms for obtaining relevant medical history, case report form (CRF) to collect treatment-related data, and adverse events form to collect data on occurrence of any adverse event during the trial.

**Fig 1.**
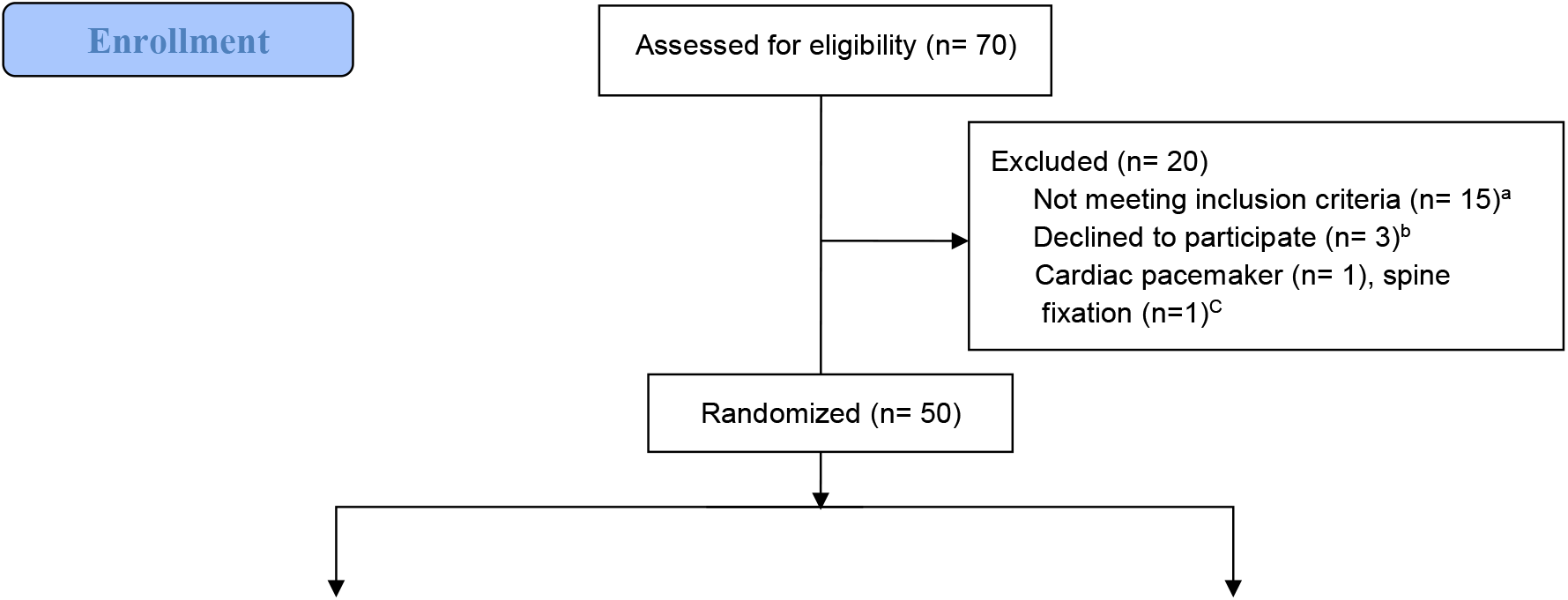
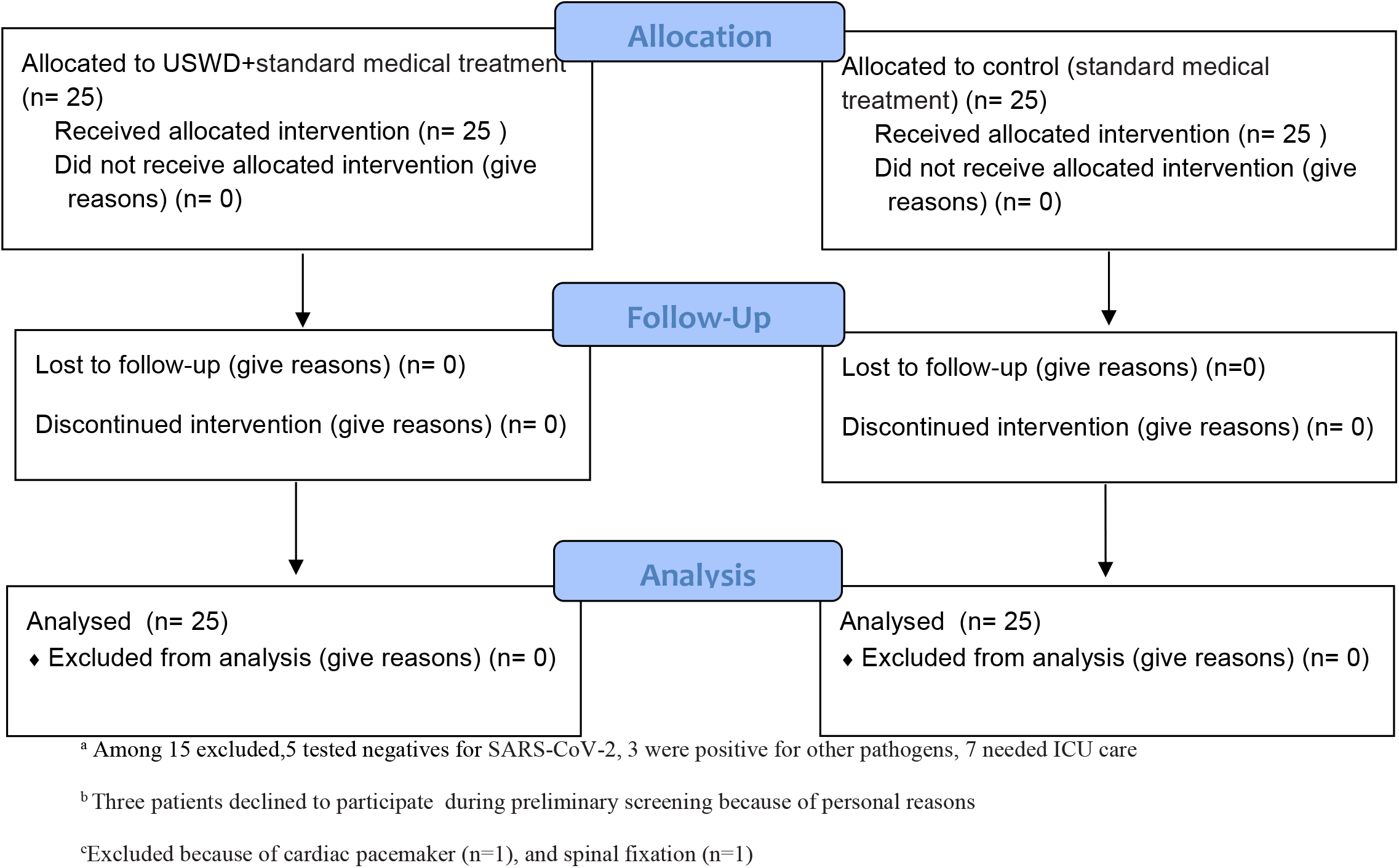
Patient Enrollment and Treatment Assignment.

### Participants

Patients of all genders qualifying the following criteria and admitted at the Tongji Hospital of Huazhong University of Science and Technology (Wuhan, China) were recruited in the study: (1) aged 18 to 65 years, (2) positive SARS-CoV-2 nucleic acid test by nasopharyngeal swabs, and (3) multiple patchy ground-glass shadows or other typical manifestations in both lungs diagnosed in chest computed tomography (CT). The exclusion criteria were: (1) positive tests for other pathogens, such as tuberculosis, mycoplasma, etc.; (2) patients with metal implants or pacemakers; (3) respiratory failure or requiring mechanical ventilation; (4) multiple organ failure requiring intensive care unit (ICU) monitoring and treatment; (5) bleeding tendency or active bleeding in the lungs; (6) shock; (7) cancer and severe underlying diseases; (8) severe cognitive impairment; (9) pregnancy or lactation; (10) those without signed informed consent; and (11) those with other contraindications to short-wave diathermy.

### Intervention

The experimental group (USWD) received the nationally recommended standard medical treatment in addition to the USWD. The USWD was performed through the application of ultra– short-wave therapy electrodes on the anterior and posterior parts of the trunk for 10 min, twice daily for 12 consecutive days. The ultra–short-wave therapy machine specifications and details are as follows: ultra–short-wave electricizer (Dajia DL-C-C, factory no: BE1003094, A.C. power 220V, 50 Hz, 700VA, Shantou Medical Equipment Factory Co., Ltd, China, Guangdong). We applied USWD in continuous mode with a frequency of 27.12MHz and a power of 200W. With these parameters, the patient would feel mild or no heat. In contrast, the control group received only the nationally recommended standard treatment. Moreover, the testing of USWD machine output, disinfection of the machine and electrodes, wearing masks, protective suits, and testing the patient’s skin sensation before the intervention were performed to ensure treatment safety.

### Outcome measures

The clinical assessment was performed at five time-points: at baseline, and after 7, 14, 21, and 28 days of treatment. The primary outcome measures were the length of recovery from symptoms measured by seven-category ordinal, systemic inflammatory response syndrome (SIRS) scale (Appendix in Supplement 1), and a negative conversion rate of the SARS-CoV-2 nucleic acid test by reverse transcription PCR (RT-PCR). The secondary outcome measures included assessment of vital signs, adverse treatment effects, CT imaging and artificial intelligence (AI)-assisted analysis, length of stay at the hospital, and blood tests, including complete blood count (CBC), creatine kinase (CK), lactate dehydrogenase (LDH), serum glutamic pyruvic transaminase (SGPT), serum glutamic oxaloacetic transaminase (SGOT), and international normalized ratio (INR). The primary and secondary variables were measured at baseline and after 7, 14, 21, and 28 days of treatment. However, CT scans were not performed frequently due to radiation hazards.

The criteria for clinical recovery were: (1) temperature returned to normal for more than 3 days; (2) significant improvement in respiratory symptoms (such as cough and breathing difficulty); (3) significant decrease in acute exudative lesions in the lung CT imaging; (4) Two consecutive negative nucleic acid test results with nasopharyngeal swabs (the sampling interval was at least 24 h).

### Date analysis

A priori sample size calculation was performed with GPower software version 3.1 (Düsseldorf, Nordrhein-Westfalen, Germany) based on the mean values of the primary variable—length of recovery from symptoms—from a previous SARS study.^17^ We estimated that with an 80% power, 5% two-sided type I error rate, and an effect size of 0.72, enrolment of 62 participants should be sufficient to detect a statistically significant between-group difference of 6.6 days in the length of recovery from symptoms. Four more participants were included in the total sample size to manage the expected 5% dropouts, making the total sample size to 66 (33 participants in each group); however, due to the subsequent unavailability of COVID-19 patients at our hospital, we had to restrict the study to 50 patients. All statistical analyses were performed using Statistical Package for Social Sciences (SPSS) version 25.0 and GraphPad Prism 8. Data normality was assessed with the Kolmogorov–Smirnov and Shapiro–Wilk tests. Continuous variables are presented as mean (standard deviation, SD) in case of normal distribution of data or median (inter-quartile range, IQR) in case of non-normal distribution, while categorical variables are presented as count (%). Descriptive statistics (mean, frequencies, and percentages) were calculated for demographic variables and primary and secondary variables in the study. Baseline and post-intervention comparisons between the USWD and control groups were performed using independent samples *t*-test and Mann–Whitney statistics based on normality results of the data. The proportions of categorical variables were compared using Fisher’s exact test/chi-square test. The Chi-square test was used for the evaluation of the seven-point scale, and the Mann–Whitney test was used for the SIRS scale (treated as ordinal scales). A difference-in-difference (D-in-D) analysis was used to analyse the AI-assisted CT scan data. Patients who failed to reach the negative conversion of SARS-CoV-2 by the cut-off date of the analysis were considered as right-censored at the last visit date. All patients were treated after completion of follow-up (28 days).

## Results

### Flow of participants through the study

Starting in February 18/2020, 70 COVID-19 patients assessed for eligibility. Among these, 20 were excluded for the reasons described in Figure 1 A total of 50 patients were randomized to either an experimental USWD group (*n* = 25) or a control group (*n* = 25). Once patients allocated in the groups, the treatment protocols started. The last clinical evaluation took place in April 20/2020

### Compliance with the trial protocol

All patients accept 12 consecutive days of USWD without interruption and completed the study protocol.There was no loss to follow-up. All participants adhered to the four follow-up sessions during 28 days

### Characteristics of the participants

Baseline characteristics are presented in Table 1. Of the 50 enrolled participants, 22 (44.0%) were men and 28 (56.0%) were women, with a mean (SD) age of 53±10.69 years. A majority of the participants were non-smokers (86.0%), and 34.0% had co-morbid conditions, such as diabetes (22%), hypertension (20%), and cardiovascular diseases (8%). Fever (90%), breathing difficulty (56%), dry cough (50%), diarrhoea (34%), and fatigue (24%) were the top five most common symptoms reported on presentation. Moreover, most of the patients had a dry cough (50%), while very few had productive cough (14% only). There were no important between-group differences in demographic data, clinical features, laboratory tests, and CT scan of the lungs at baseline(Table 2).

**Table 1.**
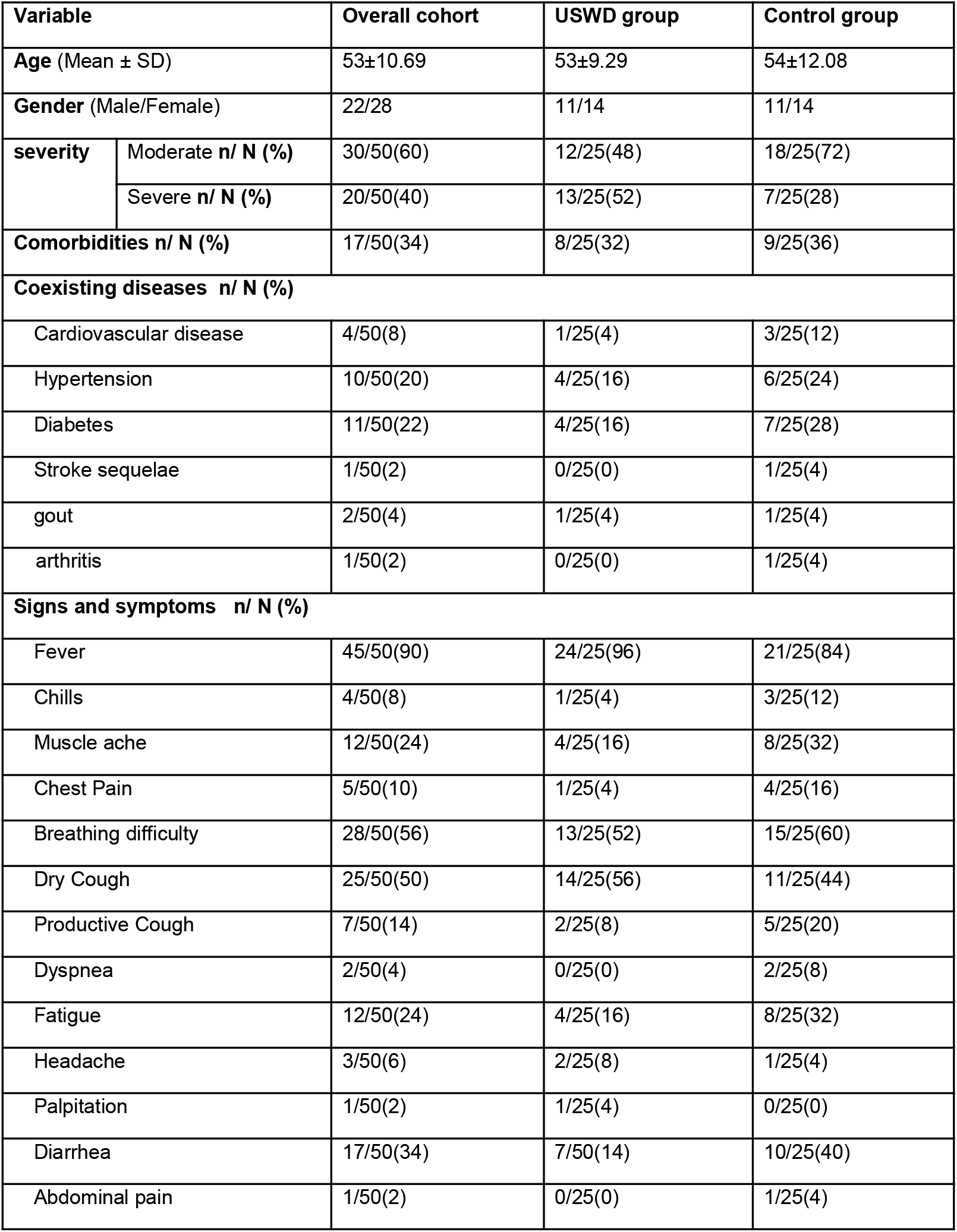
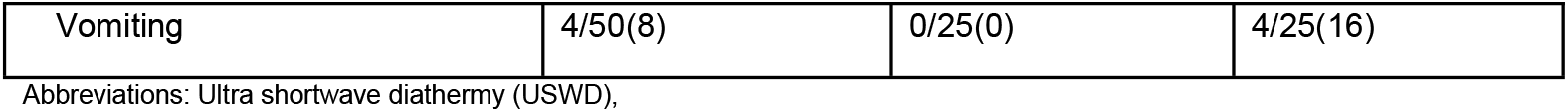
Demographics, Comorbidities, and Baseline Disease Characteristics.

| Variable |  | Overall cohort | USWD group | Control group |
| --- | --- | --- | --- | --- |
| Age (Mean ± SD) |  | 53±10.69 | 53±9.29 | 54±12.08 |
| Gender (Male/Female) |  | 22/28 | 11/14 | 11/14 |
| severity | Moderate n/ N (%) | 30/50(60) | 12/25(48) | 18/25(72) |
|  | Severe n/ N (%) | 20/50(40) | 13/25(52) | 7/25(28) |
| Comorbidities n/ N (%) |  | 17/50(34) | 8/25(32) | 9/25(36) |
| Coexisting diseases n/ N (%) |  |  |  |  |
| Cardiovascular disease |  | 4/50(8) | 1/25(4) | 3/25(12) |
| Hypertension |  | 10/50(20) | 4/25(16) | 6/25(24) |
| Diabetes |  | 11/50(22) | 4/25(16) | 7/25(28) |
| Stroke sequelae |  | 1/50(2) | 0/25(0) | 1/25(4) |
| gout |  | 2/50(4) | 1/25(4) | 1/25(4) |
| arthritis |  | 1/50(2) | 0/25(0) | 1/25(4) |
| Signs and symptoms n/ N (%) |  |  |  |  |
| Fever |  | 45/50(90) | 24/25(96) | 21/25(84) |
| Chills |  | 4/50(8) | 1/25(4) | 3/25(12) |
| Muscle ache |  | 12/50(24) | 4/25(16) | 8/25(32) |
| Chest Pain |  | 5/50(10) | 1/25(4) | 4/25(16) |
| Breathing difficulty |  | 28/50(56) | 13/25(52) | 15/25(60) |
| Dry Cough |  | 25/50(50) | 14/25(56) | 11/25(44) |
| Productive Cough |  | 7/50(14) | 2/25(8) | 5/25(20) |
| Dyspnea |  | 2/50(4) | 0/25(0) | 2/25(8) |
| Fatigue |  | 12/50(24) | 4/25(16) | 8/25(32) |
| Headache |  | 3/50(6) | 2/25(8) | 1/25(4) |
| Palpitation |  | 1/50(2) | 1/25(4) | 0/25(0) |
| Diarrhea |  | 17/50(34) | 7/50(14) | 10/25(40) |
| Abdominal pain |  | 1/50(2) | 0/25(0) | 1/25(4) |

|  |  |  |  |
| --- | --- | --- | --- |
| Vomiting | 4/50(8) | 0/25(0) | 4/25(16) |

Abbreviations: Ultra shortwave diathermy (USWD),

**Table 2.** Participants Clinical Status at Baseline.

| Variables | Overall cohort | Control group | USWD group |
| --- | --- | --- | --- |
| **Interval between onset and admission, median (IQR), d** | 21(13.25-27.00) | 18(8-35.5) | 13(8-23.0) |
| **Respiratory rate, median (IQR)** | 20.08(0.83) | 20.00(1.00) | 20.00(2.00) |
| **7-Point scale at baseline, No. (%)** |  |  |  |
| 1:Not hospitalized with the resumption of normal activities | 0(0) | 0(0) | 0(0) |
| 2:Not hospitalized, but unable to resume normal activities | 0(0) | 0(0) | 0(0) |
| 3:Hospitalized, not requiring supplemental oxygen | 4(8) | 3(12) | 1(4) |
| 4:Hospitalized, requiring supplemental oxygen | 46(92) | 22(88) | 24(96) |
| 5:Hospitalized, requiring high-flow oxygen therapy or non-invasive mechanical ventilation | 0(0) | 0(0) | 0(0) |
| 6:Hospitalized, requiring ECMO, invasive mechanical ventilation, or both | 0(0) | 0(0) | 0(0) |
| 7:Death | 0(0) | 0(0) | 0(0) |
| **SIRS scale at baseline. median (IQR)** | 2.5(1-5.0) | 3(2-5) | 2(1-4.5) |
| **Laboratory values before treatment** |  |  |  |
| Red blood cells 10<sup>9</sup>/L | 4.15(0.76) | 4.04(0.73) | 4.20(0.70) |
| White blood cells 10<sup>9</sup>/L | 6.30(2.64) | 6.68(3.88) | 5.56(2.76) |
| Neutrophil count 10<sup>9</sup>/L | 4.30(2.59) | 4.29(3.33) | 3.29(2.13) |
| Neutrophil percent % | 64.55±13.66 | 67.52±13.41 | 61.57±13.51 |
| Lymphocyte count 10<sup>9</sup>/L | 1.43(.60) | 1.43±.56 | 1.43±.56 |
| Lymphocyte percent % | 25.85(11.92) | 23.68±11.89 | 28.02±11.78 |
| Monocyte count 10<sup>9</sup>/L | 8.23±2.95 | 7.77±2.79 | 8.68±3.09 |
| ALT U/L | 32.67(22.71) | 29.00(16) | 26.00(13) |
| AST U/L | 32.22(21.40) | 23.00(14.50) | 28.00(15.50) |
| International normalised ratio ( INR ) | 2.86(12.86) | 1.05(0.10) | 1.02(0.12) |

Abbreviations: Systemic Inflammatory Response Scale (SIRS), Aspartate Aminotransferase (AST), Alanine Aminotransferase (ALT)

### Effects of the intervention

#### Primary outcomes

The time to clinical recovery (days) in the USWD group was significantly shorter than that in control group (36.84±9.93 vs. 43.56±12.15, *P* = 0.037). The SARS-CoV-2 nucleic acid test negative conversion rate showed no significant difference between the USWD and control group at days 7 (*P* = 0.066), 14 (*P* = 0.239), 21 (*P* = 0.269), and 28 (*P* = 0.490) (Table 3).

**Table 3.** Primary and Secondary Clinical Outcomes.

| Variables | USWD group<br>(n=25) | Control group<br>(n=25) | P-value |
| --- | --- | --- | --- |
| **primary clinical outcomes** |  |  |  |
| **Time to clinical recovery(Mean± SD), d** | 36.84±9.93 | 43.56±12.15 | 0.037 |
| **Viral nucleic acid negative rate, No./total (%)** |  |  |  |
| At day 7 | 2/25(8) | 7/25(28) | 0.066 |
| At day 14 | 14/25(56) | 18/25(72) | 0.239 |
| At day 21 | 22/25(88) | 19/25(76) | 0.269 |
| At day 28 | 25/25(100) | 23/25(92) | 0.490 |
| **Secondary outcomes** |  |  |  |
| Time from onset to nucleic acid negative(Mean± SD), d | 25.92±11.28 | 26.32±14.02 | 0.192 |
| **Clinical status: SIRS scale(IQR)** |  |  |  |
| At day 7 | 1 (0.5-3) | 3 (2-4) | 0.030 |
| At day 14 | 1 (0-2) | 3 (1-3.5) | 0.002 |
| At day 21 | 0 (0-1) | 2 (1-3) | 0.003 |
| At day 28 | 0 (0-1) | 1 (0-2) | 0.011 |
| **Clinical condition: 7-point scale on day 28, No. (%)** |  |  | 0.003 |
| 1:Not hospitalized with the resumption of normal activities | 19/25(76) | 7/25(28) |  |
| 2:Not hospitalized, but unable to resume normal activities | 3/25(12) | 13/25(52) |  |
| 3:Hospitalized, not requiring supplemental oxygen | 3/25(12) | 3/25(12) |  |
| 4:Hospitalized, requiring supplemental oxygen | 0/25(0) | 2/25(8) |  |
| 5:Hospitalized, requiring high-flow oxygen therapy or non-invasive mechanical ventilation | 0/25(0) | 0/25(0) |  |
| 6:Hospitalized, requiring ECMO, invasive mechanical ventilation, or both | 0/25(0) | 0/25(0) |  |
| 7:Death | 0/25(0) | 0/25(0) |  |
| **Laboratory values after treatment** |  |  |  |
| Red blood cells 10<sup>9</sup>/L | 4.05(0.80) | 4.03(0.62) | 0.62 |
| white blood cells 10<sup>9</sup>/L | 5.51±1.38 | 6.56±1.97 | 0.035 |
| Neutrophil count 10<sup>9</sup>/L | 2.99(1.49) | 3.83(1.59) | 0.071 |
| Neutrophil percent | 58.49±7.24 | 58.55±9.37 | 0.980 |
| Lymphocyte count 10<sup>9</sup>/L | 1.60(0.63) | 1.79(0.97) | 0.207 |
| Lymphocyte percent | 29.98±6.96 | 28.79±8.42 | 0.589 |
| Monocyte count 10<sup>9</sup>/L | 8.92(2.20) | 7.10(1.15) | 0.002 |
| ALT U/L | 23.00(13.0) | 24.00(16.0) | 0.771 |
| AST U/L | 23.00(7.0) | 19.00(10.0) | 0.264 |
| international normalized ratio (INR ) | 1.03(0.09) | 1.00(0.06) | 0.022 |
| **Complications n/ N(%)** |  |  |  |
| Abnormal LFTs | 13/25(52) | 12/25(48) | 0.777 |
| Elevated AST | 7/25(28) | 7/25(28) | 1.000 |
| Elevated ALT | 12/25(48) | 12/25(48) | 1.000 |
| Electrolyte imbalance | 8/25(32) | 11/25(44) | 0.382 |
| Hyperfibrinogenemia | 11/25(44) | 12/25(48) | 0.777 |
| Anemia | 8/25(32) | 13/25(52) | 0.152 |
| Hypoalbuminemia | 11/25(44) | 6/25(24) | 0.136 |
| Abnormal blood coagulation | 2/25(8) | 2/25(8) | 1.000 |
| Renal insufficiency | 4/25(16) | 1/25(4) | 0.349 |
| Myocardial damage | 1/25(4) | 1/25(4) | 1.000 |
| No complications | 3/25(12) | 3/25(12) | 1.000 |

Abbreviations: Ultra shortwave diathermy (USWD), Systemic Inflammatory Response Scale (SIRS), Aspartate Aminotransferase (AST), Alanine Aminotransferase (ALT), liver function tests (LFTs).

#### Secondary outcomes

##### Clinical outcomes

The SIRS scores, which reflect patients’ present clinical condition (including heart and respiratory rate, mean arterial pressure, SpO_2_ %, body temperature, white blood cells, and level of consciousness), were statistically different between the two groups at days 7 (*P* = 0.030), 14 (*P* = 0.002), 21 (*P* = 0.003), and 28 (*P* = 0.011). Moreover, the seven-point scale after intervention at days 21 and 28 also showed significant differences between the two groups (*P* = 0.002, and *P* = 0.003, respectively); however, the difference at day 7 and 28 was statistically insignificant (*P* = 0.524, *P* = 0.108). These findings suggest the therapeutic efficacy of implementing USWD in patients with COVID-19 (Table 3).

#### CT scan outcomes

In Figures 2 and 3, the CT images depict the treatment progress in moderate and severe cases in both the groups throughout the interventional period. An AI-aided CT (AI-aided CT) image analysis system was adopted for the quantitative analysis of the infected lung area proportion and volume before and after treatment. The mean values of different CT scan parameters for the total population before and after treatment are shown in Table 4. The lower lung had the worst infective conditions; and in CT, both the groups showed improvement in the infected lung area proportion and volume. A comparison of AI-assisted quantitative analysis of CT scan images before and after treatment showed no significant differences between the control and USWD groups (Table 5 and Figure 3).

**Table 4.**
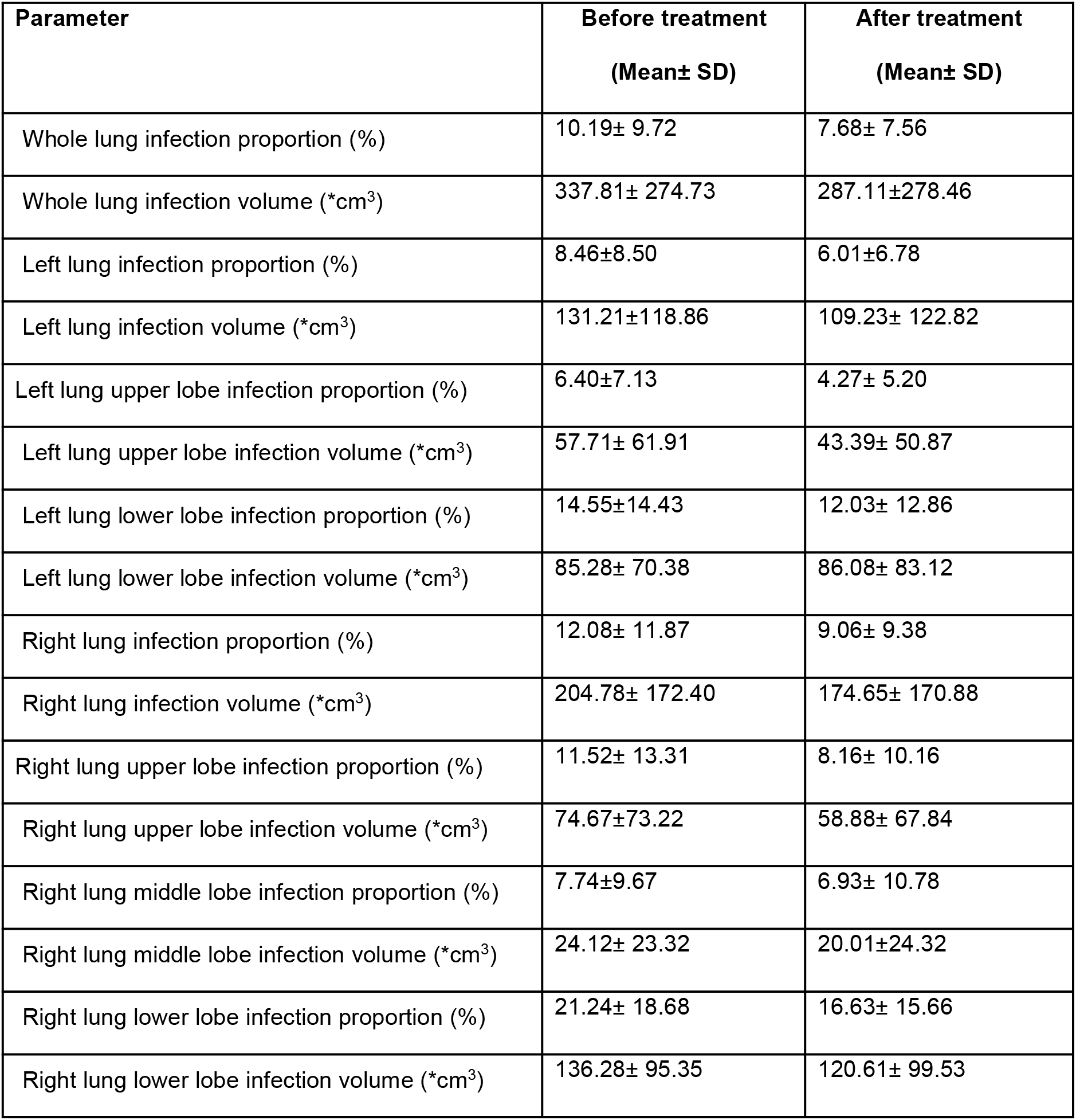
AI-assisted CT scan parameters of the total population before and after treatment.

**Table 5.** Comparison of Mean AI-assisted CT scan parameters between USWD and control group.

| Variables | 0group |  |  | 1group |  |  | D-in-D | P |
| --- | --- | --- | --- | --- | --- | --- | --- | --- |
|  | Before | After | Change<sup>a</sup> | Before | After | Change<sup>a</sup> |  |  |
| whole lung infection proportion% | 8.2(8.1) | 6.8(8.1) | -1.4 | 12.8(10.8) | 8.7(7.5) | -3.8 | -2.29 | 0.5243 |
| whole lung infection V cm<sup>3</sup> | 297.1 (268.7) | 250.9 (256.6) | -46.2 | 395.0 (274.5) | 325.3 (312.0) | -69.7 | -0.163 | 0.9988 |
| Lt lung infection proportion | 6.8 (7.5) | 5.6 (7.0) | -1.2 | 10.5 (9.2) | 6.5 (7.0) | -4 | -2.38 | 0.4607 |
| Lt lung infection V cm<sup>3</sup> | 114.2(118.9) | 100.2(111.1) | -14 | 154.7(117.1) | 119.6(141.5) | -35.1 | -12.45 | 0.7995 |
| Lt lung upper lobe infection proportion | 5.1(6.6) | 4.0(6.4) | -1.1 | 6.6(7.5) | 3.5(3.8) | -3.1 | -1.73 | 0.5107 |
| Lt lung upper lobe infection V cm<sup>3</sup> | 49.4(69.8) | 37.9(56.7) | -11.5 | 56.0(52.9) | 38.0(45.0) | -18 | -3.72 | 0.8748 |
| Lt lung lower lobe infection proportion | 10.1(12.2) | 8.8(9.8) | -1.3 | 17.3(15.8) | 12.2(15.7) | -5.1 | -3.07 | 0.5839 |
| Lt lung lower lobe infection V cm<sup>3</sup> | 61.7(59.4) | 66.5(60.8) | 4.8 | 98.7(78.7) | 83.3(104.6) | -15.4 | -14.53 | 0.6399 |
| Rt lung infection proportion | 9.9(10.0) | 7.8(9.4) | -2.1 | 14.9(13.3) | 10.5(9.9) | -4.4 | -1.61 | 0.7158 |
| Rt lung infection V cm<sup>3</sup> | 182.9(162.9) | 148.8(155.1) | -34.1 | 236.5(179.7) | 205.3 (191.5) | -31.2 | 17.60 | 0.7946 |
| Rt lung upper lobe infection proportion | 8.5(10.8) | 6.3(9.1) | -2.2 | 11.0(14.9) | 7.8(11.1) | -3.2 | -0.41 | 0.9332 |
| Rt lung upper lobe infection V cm<sup>3</sup> | 58.0(66.7) | 45.0(60.7) | -13 | 68.6(79.1) | 57.2(74.8) | -11.4 | 7.46 | 0.7945 |
| Rt lung middle lobe infection proportion | 4.4(6.4) | 4.2(7.2) | -0.2 | 9.2(11.4) | 7.8(12.9) | -1.4 | -0.67 | 0.8691 |
| Rt lung middle lobe infection V cm<sup>3</sup> | 16.1(17.6) | 15.2(20.6) | -0.9 | 26.9(27.1) | 19.6(27.4) | -7.3 | -4.42 | 0.6391 |
| Rt lung lower lobe infection proportion | 15.8(17.5) | 12.4(16.0) | -3.4 | 24.1(19.6) | 17.7(16.0) | -6.4 | -2.00 | 0.7769 |
| Rt lung lower lobe infection V cm<sup>3</sup> | 110.6(100.2) | 88.3(91.5) | -22.3 | 145.3(94.8) | 130.6(111.5) | -14.7 | 16.93 | 0.6594 |

<sup>a</sup> Average change in means before and after treatment, abbreviations: D-in-D=Difference-in-difference), V=volume, Rt=right, Lt=left

**Fig 2.**
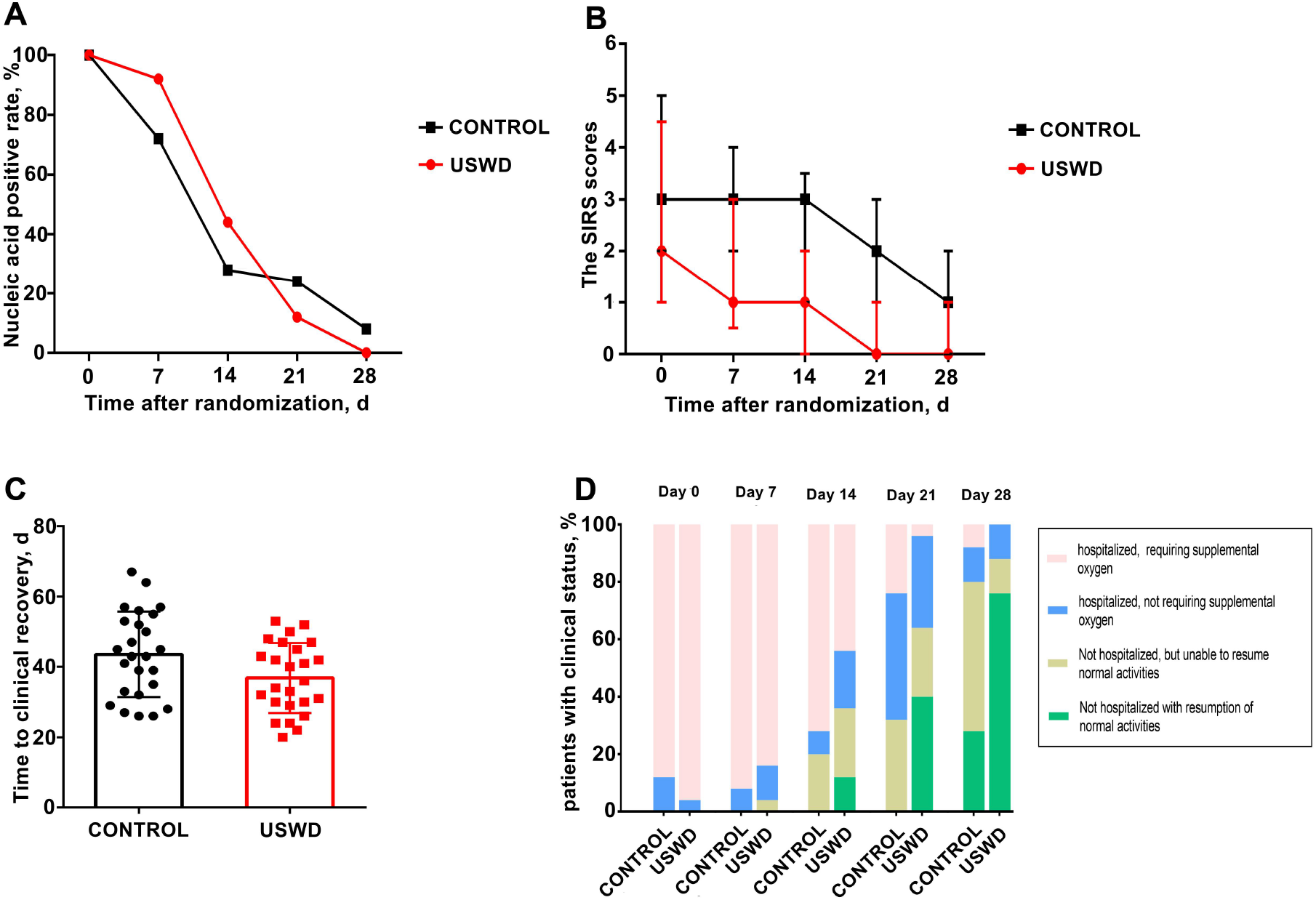
the outcomes on days 7, 14, 21 and 28 by treatment group. (A) The SARS-CoV-2 nucleic acid negative conversion rate showed no significant difference between the USWD and control group at day 7 (P = 0.066), day14 (P = 0.239), day 21 (P = 0.269), and day 28 (P = 0.490). (B) The clinical condition on SIRS score showed statistically significant difference on day 7 (P = 0.030), day14 (P = 0.002), day 21 (P = 0.003) and day 28(P = 0.011). C Time to clinical recovery in the USWD group was significantly shortened comparing with control group (P = 0.037). D Clinical status on 7-point ordinal scale on study days 21 and 28 showed significance (P = 0.002, 0.003), whereas the difference at day 7 and 14 was insignificant (P = 0.524, 0.108).

**Figure 3.**
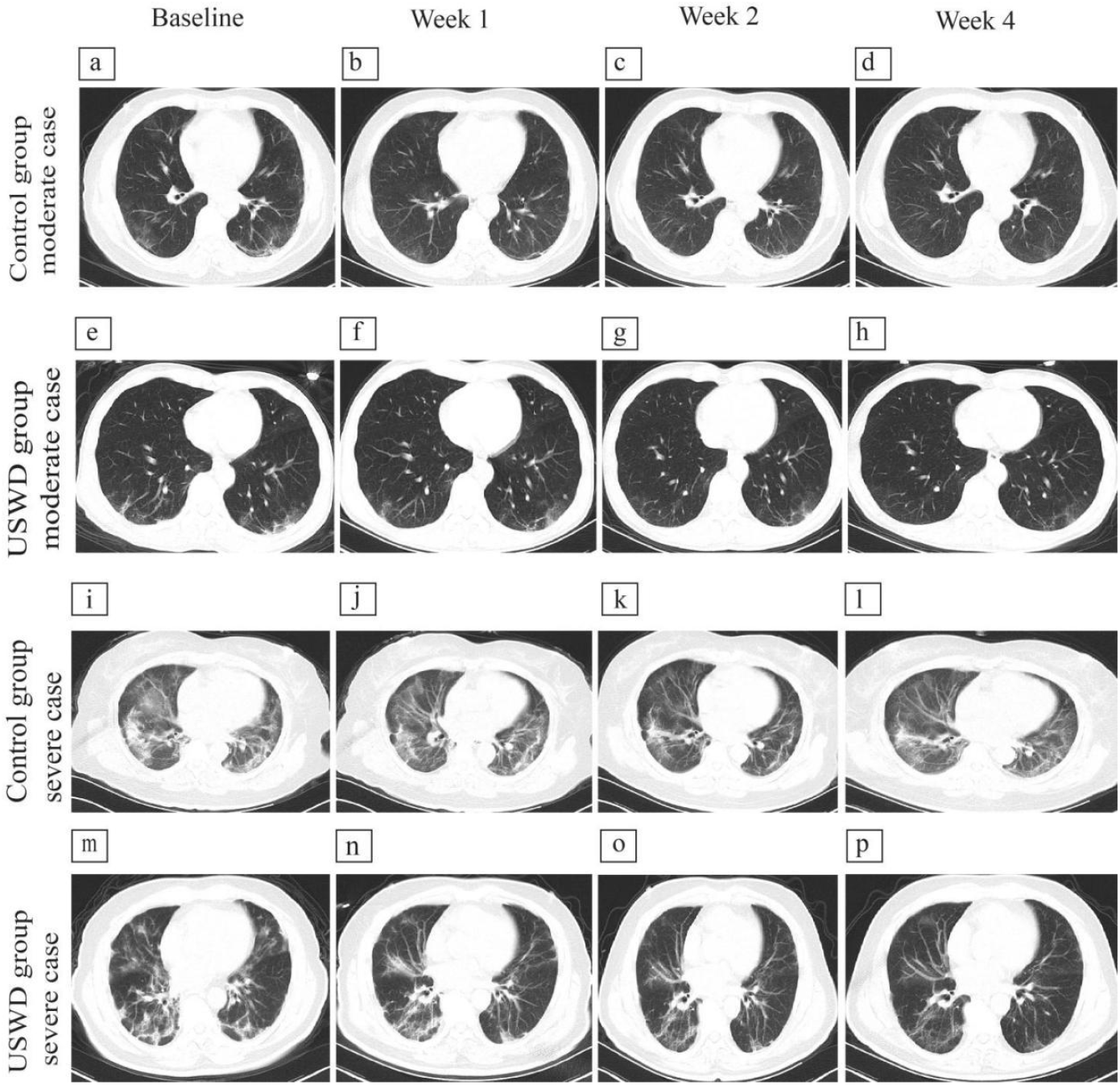
Chest CT images of moderate and severe cases in control and ultra–short-wave diathermy (USWD) groups. **(a-d)** Moderate cases in control group. Multiple ground glass opacity (GGO) in both lower lungs, with local thickening and adhesion of bilateral pleura at baseline, while GGO significantly absorbed finally. Moreover, streak shadows are also lighter, and pleural thickening and adhesions alleviated at week-4. No lung consolidation, pulmonary fibrosis, pleural effusion, or pulmonary interstitial changes found. **(e-h)** Moderate cases in USWD group. Bilateral scattered GGO in the lower lungs, with grid shadows and striae foci visible inside, obvious near the pleura, and local pleural adhesions are observed at baseline, while bilateral scattered GGO appear much lightened than before with a significant decrease at week-4. Bilateral local pleural slight adhesion. No obvious lung consolidation and pulmonary fibrosis are observed. **(i-l)**: Severe cases in control group. Bilateral multiple GGO, accompanied by consolidation shadows and a few cord lesions, which are obvious near the pleura. Thickening and adhesion of bilateral pleura, while GGO still existed with slight alleviation. No obvious changes in consolidation shadows, grid shadows, and cord lesions. Thickening, and adhesion of bilateral pleura are visible, with some pulmonary fibrosis. **(m-p)** Severe cases in USWD group. Bilateral multiple GGO, and striped shadows, accompanied by bronchial inflation, and local thickening and adhesion of the pleura on both sides at baseline, while alleviated at week-4. Moreover, the bronchial inflation disappeared, and grid shadows and fibrous cord shadows improved. Both sides of the pleura are slightly thickened and adherent. A little pulmonary fibrosis and local mild pulmonary consolidation are visible with no pleural effusion.

#### Adverse events and complications

No serious AEs, deaths, permanent disability, neoplasia, or empyrosis cases were registered during the trial. In the USWD group, 14/15 cases of pulmonary fibrosis recovered, one had no change in fibrosis; similarly, in the control group, 16/18 cases of fibrosis recovered, and two had no changes in fibrosis. Worsening of pulmonary fibrosis was observed in neither group. Out of 50 patients, 22 each in the USWD and control groups had complications, such as abnormal liver function test (LFT; 52% vs. 48%, *P* = 0.777), electrolyte imbalance (32% vs. 44%, *P* = 0.382), hyperfibrinogenaemia (44% vs. 48%, *P* = 0.777), and mild anaemia (32% vs. 52%, *P* = 0.152). Routine blood investigation showed all parameters in almost equal and normal ranges in both the groups. However, the WBC counts were significantly lower in the USWD group than in control group (5.51±1.38 vs. 6.56±1.97). In contrast, the median monocyte count was significantly higher in USWD than in control group (8.92 [2.20] vs. 7.10 [1.15]), but the difference was of uncertain clinical importance.

## Discussion

To the best of our knowledge, this is the first randomized clinical trial investigating the efficacy of USWD treatment in COVID-19 (a search of PubMed and MEDLINE on 31 October 2020 for publications in all languages using the keyword ‘COVID-19’ revealed no published USWD randomized clinical trials). In this randomized clinical trial, we systematically investigated the safety and therapeutic efficacy of USWD in patients with moderate or severe COVID-19. There was a significant shortening in the length of recovery and improvement in the mean scores of the clinical scales, including SIRS and the seven-point scale after a 12-days course of USWD administered twice daily.

The USWD is known to enhance fibroblast activity.^18^ Fibroblasts are oxygen-sensitive cells,^19^ and theoretically, the synergistic activity of USWD and high oxygen environment in COVID-19 patients could cause or aggravate pulmonary fibrosis. Interestingly, worsening of pulmonary fibrosis was not observed in patients in this study. In contrast, fibrosis was alleviated in 14 out of 15 patients in the USWD group and 16 out of 18 patients in control group after the treatment.

In this study, the SARS-CoV-2 negative conversion rate was not enhanced by USWD, suggesting that USWD exerts therapeutic function independent of the direct antiviral effect. The USWD generates radiations of 27.12 MHz. When applied in a continuous mode, USWD can induce vasodilation, enhance cellular activity, and reduce inflammation and pain in the patients. The findings of this study are consistent with those of Zhang et al.; they used USWD in 2003 during the SARS for treating 38 patients, which conferred significant improvements in the patients when used as an adjuvant treatment with drug therapy, and the administration of USWD accelerated patient recovery and shortened the duration of hospital stay.^17^ Many other studies with pneumonia patients treated with USWD show similar results about clinical recovery as those in our study: He YG 2006^20^ administered USWD in children with bronchopneumonia, which reduced inflammation, enhanced lung tissue repair, and immune response; Du QP 2012^21^ combined USWD with other therapies in infants with pneumonia and reported that adjuvant USWD reduced the time of disappearance of symptoms, shortened the treatment course, and reduced the use of antibiotics; Zhu Q 1999^22^ reported that combined administration of medications and USWD can impart better effects on pulmonary function and clinical recovery signs.

Treatment with USWD, however, increased the number of monocytes, which is an important component of the body’s immune system, although within normal range; and reduced the number of WBCs, which is a marker of inflammation. These findings are consistent with those in previous studies of the physiological effects of short wave therapy, supporting the immune response to accelerate recovery. Moreover, the findings in this study concur with those of Bazett et al., suggesting that USWD could significantly increase leukocytes ability to perform phagocytosis and adhere to the vessel walls; thus, administration of USWD at an early stage in pneumonia may stimulate and boost body’s natural defences against microorganisms.^23,24^ Lung CT scans provide supportive assistance in the early diagnosis and recovery monitoring of lung lesions in patients with COVID-19. Additionally, AI-aided CT analysis can be used to scientifically identify the severity and quantification of lung lesions in patients with COVID-19.^25,26^

We attempted AI-aided CT analysis to compare the treatment effects on lung involvement in patients between the two groups in our study. The CT quantitative analysis showed no significant differences in the USWD and control groups; however, in most of the patients, the fibrosis observed before treatment was recovered (recovery: USWD=14/15 and control=16/18). The fibrosis recovery result could completely overcome the safety concerns about USWD. Moreover, administration of USWD improved condition in patients with COVID-19 who were hospitalized and required supplemental oxygen therapy. However, due to early termination of the study and the small sample size, the implications of the findings of this study were limited.

This study had some strengths and limitations. This is the first randomized controlled trial addressing the safety and efficacy of USWD in patients with COVID-19 admitted at the Tongji Hospital, China, which is compliant with good clinical practices. However, this study had some limitations, including early termination, small sample size, and a single-centre design.

In conclusion, Administration of USWD as an adjunctive therapy to standard therapy shortens the disease course in patients with moderate and severe COVID-19 without aggravating pulmonary fibrosis. Based on the beneficial clinical effects and no adverse effects of USWD, we recommend the use of USWD as an adjunct to standard therapies in patients with COVID-19.

Nevertheless, we suggest studies with a larger sample size to confirm these findings and further explore the benefits of USWD.

### Clinical recommendations

- USWD as an adjunctive therapy shortened the recovery course and improved the clinical status of patients with COVID-19 without significant adverse events.
- USWD group showed no improvement in the negative SARS-CoV-2 conversion rate, suggesting that the therapeutic effect of USWD does not depend on direct antiviral activity.

## Data Availability

The trial data could be provided by a reasonable request to the corresponding authors (HC).

http://www.chictr.org.cn/historyversionpuben.aspx?regno=ChiCTR2000029972

## Supplement 1 Appendix

### Systemic Inflammatory Response scale (SIRS)

Systemic Inflammatory Response scale (SIRS) is used for evaluation of clinical improvement based on heart rate, mean arterial pressure(MAP mmHg), respiratory rate/min, blood oxygen saturation (SpO_2_ %), body temperature °C), white blood cells (WBC *10^9^/L), blood glucose (mmol/L) and level of consciousness (Aware/awake, Lethargy or irritability, shallow coma, coma, brain death). All these parameters, except” Level of consciousness” are assigned a score from 0 to 4 based on the actual values recorded from the patient corresponding to the range of values in the table below. For the level of consciousness: Aware/awake (0), Lethargy or irritability (1), Shallow coma (2), coma (3), brain death (4).

### Systemic Inflammatory Response scale (SIRS)

| Description | 0 points | 1 points | 2 points | 3 points | 4 points | score |
| --- | --- | --- | --- | --- | --- | --- |
| Heart beat | 60-100 | 55-59;<br>100-119 | 50-54;<br>120-140 | 41-49;<br>141-160 | <40;<br>>160 |  |
| MAP(mmHg) | 70-100 | 60-69;<br>101-110 | 50-59;<br>111-130 | 40-49;<br>131-159 | <40;<br>>160 |  |
| Respiratory rate /<br>min | 12-20 | 9-12;<br>20-25 | 5-8;<br>26-35 | <5;<br>36-45 | 0;<br>>46 |  |
| SpO<sub>2</sub> ( % ) | >92 | 85-91 | 75-84 | 60-74 | <60 |  |
| body temperature<br>( °C ) | 36.0-37.5 | 35-35.9;<br>37.5-38.5 | 34-34.9;<br>38.6-39.5 | 33-33.9;<br>39.6-40 | <33;<br>>40 |  |
| WBC(*10<sup>9</sup>/L) | 4.0-10.0 | 3.0-3.9;<br>10.1-14.9 | 2.0-2.9;<br>15-20.0 | 1.0-2.0;<br>20.1-30.0 | <1.0;<br>>30.1 |  |
| GLU(mmol/L) | 3.5-5.5 | 5.7-8.6 | 8.7-13.5 | 13.6-23 | >23 |  |
| Level of<br>consciousness | Aware/awake | Lethargy or<br>irritability | Shallow coma | coma | brain death |  |
| **Total score :** |  |  |  |  |  |  |

### The 7-category ordinal

The 7-category or 7-point ordinal scale is consisting of seven separate categories. The categories range from 1-7, 7 corresponds to death; category 6 patients need ICU hospitalization, require ECMO and invasive mechanical ventilation; category 5 patients need ICU hospitalization but do not require ECMO and/or invasive mechanical ventilation; category 4 patients are non-ICU hospitalized patients, requiring supplemental oxygen; category 3 patients are also non-ICU hospitalized patients, but they do not require supplemental oxygen; category 2 patients are not hospitalized, but unable to resume normal activities; category 1, not hospitalized with the resumption of normal activities.

## References

1. Epidemiology Working Group for NCIP Epidemic Response, Chinese Center for Disease Control and Prevention. The epidemiological characteristics of an outbreak of 2019 novel coronavirus diseases (COVID-19) in China. Zhonghua Liu Xing Bing Xue Za Zhi. 2020;41(2):145–151. doi: 10.3760/cma.j.issn.0254-6450.2020.02.003.

2. Cucinotta D, Cucinotta D, Vanelli M. WHO Declares COVID-19 a Pandemic. Acta Biomed. 2020;91(1):157–160. doi: 10.23750/abm.v91i1.9397.

3. Bedford J, Enria D, Giesecke J, et al. COVID-19: towards controlling of a pandemic. Lancet. 2020;395(10229):1015–1018. doi: 10.1016/S0140-6736(20)30673-5.

4. Peeri NC, Shrestha N, Rahman MS, et al. The SARS, MERS and novel coronavirus (COVID-19) epidemics, the newest and biggest global health threats: what lessons have we learned? Int J Epidemiol. 2020;49(3):717–726. doi: 10.1093/ije/dyaa033.

5. Strzelecki A. The second worldwide wave of interest in coronavirus since the COVID-19 outbreaks in South Korea, Italy and Iran: A Google Trends study. Brain Behav Immun. 2020;88:950–951. doi: 10.1016/j.bbi.2020.04.042.

6. Yang LL, Yang T. Pulmonary rehabilitation for patients with coronavirus disease 2019 (COVID-19). Chronic Dis Transl Med. 2020;6(2):79–86. doi: 10.1016/j.cdtm.2020.05.002.

7. Thomas P, Baldwin C, Bissett B, et al. Physiotherapy management for COVID-19 in the acute hospital setting: clinical practice recommendations. J Physiother. 2020;66(2):73–82. doi: 10.1016/j.jphys.2020.03.011.

8. Lazzeri M, Lanza A, Bellini R, et al. Respiratory physiotherapy in patients with COVID-19 infection in acute setting: a Position Paper of the Italian Association of Respiratory Physiotherapists (ARIR). Monaldi Arch Chest Dis. 2020;90(1). doi: 10.4081/monaldi.2020.1285.

9. Dantas LO, Barreto R, Ferreira C. Digital physical therapy in the COVID-19 pandemic. Braz J Phys Ther. 2020;24(5):381–383. doi: 10.1016/j.bjpt.2020.04.006.

10. Yu HP, Jones AY, Dean E, et al. Ultra-shortwave diathermy - a new purported treatment for management of patients with COVID-19. Physiother Theory Pract. 2020;36(5):559–563. doi: 10.1080/09593985.2020.1757264.

11. Lamina S, Hanif S, Gagarawa YS. Short wave diathermy in the symptomatic management of chronic pelvic inflammatory disease pain: A randomized controlled trial. Physiother Res Int. 2011;16(1):50–6. doi: 10.1002/pri.473.

12. Vardiman JP, Moodie N, Siedlik JA, et al. Short-Wave Diathermy Pretreatment and Inflammatory Myokine Response After High-Intensity Eccentric Exercise. J Athl Train. 2015;50(6):612–20. doi: 10.4085/1062-6050-50.1.12.

13. Goats GC. Continuous short-wave (radio-frequency) diathermy. Br J Sports Med. 1989;23(2):123–7. doi: 10.1136/bjsm.23.2.123.

14. Laufer Y, Zilberman R, Porat R, et al. Effect of pulsed short-wave diathermy on pain and function of subjects with osteoarthritis of the knee: a placebo-controlled double-blind clinical trial. Clin Rehabil. 2005;19(3):255–63. doi: 10.1191/0269215505cr864oa.

15. Shields N, Gormley J, O’Hare N. Short-wave diathermy: current clinical and safety practices. Physiother Res Int. 2002;7(4):191–202. doi: 10.1002/pri.259.

16. Gorbunov FE, Vinnikov AA, Strelkova NI, et al. [The use of pulsed and continuous UHF electrical fields in the rehabilitation of patients with the Guillain-Barre syndrome and other peripheral myelinopathies]. Zh Nevrol Psikhiatr Im S S Korsakova. 1995;95(5):22–6.

17. Zhang L, Zheng G, Liu G, et al. Application of ultrashort wave diathermy in treatment of severe acute respiratory syndrome. Chinese J Rehabil Med. 2003;25(6):332–334. doi:10.3760/j:issn:0254-1424.2003.06.005.

18. Hill J, Lewis M, Mills P, et al. Pulsed short-wave diathermy effects on human fibroblast proliferation. Arch Phys Med Rehabil. 2002;83(6):832–6. doi: 10.1053/apmr.2002.32823.

19. Hu Y, Fu J, Xue X. Association of the proliferation of lung fibroblasts with the ERK1/2 signaling pathway in neonatal rats with hyperoxia-induced lung fibrosis. Exp Ther Med. 2019;17(1):701–708. doi: 10.3892/etm.2018.6999.

20. He Y, Ruan Q, Chang X, et al. Changes of serum cytokines in children with bronchopneumonia treated with ultrashort wave diathermy. J Appl Clin Pediatr. 2006;21(4):220. doi:10.3969/j.issn.1003-515X.2006.04.015.

21. Du Q, Nan J, Qin L. Benefits of adjuvant therapy with transcutaneous ion introduction of traditional Chinese medicine by using microcomputer control intermediate frequency stimulator and ultrashort wave therapy on 112 cases of infantile pneumonia. Chinese J Gen Pract. 2012;10(8):1232-1233. doi:10.16766/j.cnki.issn.1674-4152.2012.08.063.

22. Zhu Q, Sun Y, Zhang A. Effect of ultrashort wave and medications on pulmonary function of pneumonia patients. CHINESE J Phys Ther. 1999;(2):2. doi:10.1016/B978-008043005-8/50012-3.

23. Coulter JS. Medical diathermy. JAMA. 1936;106(3):209–214. doi:10.1001/jama.1936.02770030004053.

24. Bazett, H. C., Mock, H. E., Pemberton, R., et al. Principles and Practice of Physical Therapy, Hagerstown, Md., W. F. Prior Company, Inc.1935, vol. 1, p. 29.

25. Vardhanabhuti V. CT scan AI-aided triage for patients with COVID-19 in China. Lancet Digit Health. 2020;2(10):e494–e495. doi: 10.1016/S2589-7500(20)30222-3.

26. Wang M, Xia C, Huang L, et al. Deep learning-based triage and analysis of lesion burden for COVID-19: a retrospective study with external validation. Lancet Digit Health. 2020;2(10):e506–e515. doi: 10.1016/S2589-7500(20)30199-0.

